# Spatial and Temporal Distribution of Visceral Leishmaniasis in Karamoja Region, Uganda: Analysis of surveillance data, 2015–2022

**DOI:** 10.1101/2024.02.16.24302916

**Authors:** Mercy Wendy Wanyana, Richard Migisha, Patrick King, Benon Kwesiga, Daniel Kadobera, Lilian Bulage, Alex Riolexus Ario

## Abstract

**Background:** Uganda targets to control leishmaniasis and eliminate visceral leishmaniasis as a public health problem by 2030, with 85% of the cases detected, 95% treated, and visceral leishmaniasis eliminated (<1% case fatality rate [CFR]). However, little is documented on the country’s progress towards achieving these targets. We examined the temporal trends and spatial distribution of leishmaniasis in the endemic Karamoja Region of Uganda, 2015–2022.

**Methods:** We analysed aggregate secondary data on clinically diagnosed leishmaniasis laboratory-confirmed cases, visceral leishmaniasis cases, hospital admissions, and deaths from the District Health Information System 2. We used population-based calculations to determine the annual prevalence of leishmaniasis and quarterly prevalence of visceral leishmaniasis per 1,000,000 persons, while the prevalence of leishmaniasis admissions and CFRs were calculated per 100 cases. We used the Mann-Kendall test to assess the significance of the trend.

**Results:** Overall, 4,008 cases of clinically diagnosed leishmaniasis were reported, and of these, 11% were laboratory-confirmed. The average annual prevalence of leishmaniasis was 4 per 1,000,000 population. From 2020 to 2022, there was an increasing trend in quarterly prevalence of visceral leishmaniasis (Kendall’s score=36, p=0.016), averaging 3 cases per 1,000,000 population. Leishmaniasis admissions increased annually to 55 per 100 cases (Kendall’s score=23, p=0.006). The average annual CFR was 5%, with no deaths reported in 2018, 2019, and 2021. Amudat District had the highest prevalence rates of leishmaniasis (477 per 1,000,000 population) and visceral leishmaniasis (139 per 100,000 population).

**Conclusion:** The increasing trend of visceral leishmaniasis, and CFR >1% threaten the goal of controlling leishmaniasis as a public health problem by 2030. Gaps in case detection may further prevent the achievement of targets. Strengthening existing interventions, such as vector control and rapid diagnostic kits for early detection and treatment, may be crucial to sustain progress toward elimination targets.

## Background

Leishmaniasis is a vector-borne neglected tropical disease caused by a protozoan and transmitted by a female sand fly mainly of *Phlebotomus* species^1^. Transmission occurs when an infected female sand fly bites a human host and transmits the parasite to the host during its blood meal^2^. The disease clinically manifests in three forms: cutaneous, mucocutaneous and visceral^3^. Clinical presentation usually depends on the virulence of the parasite, the individual’s immunity, and the site of the lesion^4^. The cutaneous form is the mildest form, typically presenting with painless nodules, macules, papules or ulcerative lesions on the skin in areas exposed to bites of the sand fly. It is commonly linked to the *Leishamania major* and *Leishmania tropica species*^2^. The mucosal form typically presents with lesions on the mucus membranes and cartilage in the nose, mouth, upper respiratory tract, and pharynx^5^. Visceral leishmaniasis manifests as fever, weight loss, splenomegaly, hepatomegaly, and reduced blood cell production, potentially causing anemia, bleeding, and susceptibility to other infections^6,7^ Visceral Leishmaniasis, often associated with L. donovani in East Africa, is highly fatal with a 95% untreated case fatality rate and 10-20% with treatment^8,9^. It carries outbreak potential, with sequelae like relapse and post-kala-azar dermal leishmaniasis, contributing to interepidemic reservoirs and sustaining transmission.

As of 2019, approximately 4,580,000 cases of leishmaniasis were reported globally, causing an estimated 657,000 new cases annually and resulting in 697,000 Disease Adjusted Life Years (DALYs) lost^10^. African countries including Uganda contribute a significant proportion of the burden, accounting for 34-57% ^11^. Leishmaniasis disproportionately impacts the poorest populations due to factors such as inadequate housing, poor environmental sanitation, lack of control measures, and economically driven migration to vector-prone areas, increasing their susceptibility to the disease ^12,13^. Children and immunocompromised individuals, including People Living with HIV (PLWHIV), are more affected than adults and non-immunocompromised individuals^14,15^.

Leishmaniasis places a catastrophic financial burden on affected households with a median cost ranging from USD 165 to 1,500 per episode^13,16^. To avert this burden, current global efforts aim at controlling leishmaniasis and eliminate leishmaniasis by 2030 through the World Health Organization(WHO) Roadmap for neglected tropical diseases 2021–2030^17^. Specifically, this aims to ensure that 85% of all leishmaniasis cases are detected, 95% treated, and visceral leishmaniasis eliminated (<1% case fatality rate [CFR]); this includes Uganda where the disease is endemic especially in the Karamoja Region. Currently there is a paucity of data on the country’s progress and burden of leishmaniasis. This could be attributed to the limited analysis of surveillance data despite it’s being in the national integrated disease surveillance system.^18^ Analysis of the surveillance data is key to detect remaining or re-emerging hotspots of leishmaniasis transmission which supports timely adaptation of control and elimination interventions^19^. Given the complexity of leishmaniasis management and diagnostics, control efforts adopt an innovative and intensified disease management approach, emphasizing individual care at health facilities^20^. As such, understanding trends in health facility leishmaniasis data is crucial for evidence-based decision-making. We examined the temporal trends and spatial distribution of leishmaniasis in the endemic Karamoja Region of Uganda, 2015–2022.

## Methods Study settings

We utilised leishmaniasis surveillance data generated from all health facilities in the Karamoja region in the years 2015–2022. Leishmaniasis is endemic in the Karamoja region in Uganda. The semi-arid climate and termite hills in this region provide a favourable environment for sand flies of *Phlebotomus* species which are the vector responsible for transmission of Visceral Leishmaniasis^21^. Within Uganda health facilities leishmaniasis is diagnosed clinically and confirmed using a Giemsa-stain slides viewed under a microscope.

### Study design and data Source

We conducted a descriptive study using routinely collected leishmaniasis surveillance data collected in District Health Information System 2 (DHIS-2) .The DHIS-2 collects data on diseases of public health importance like leishmaniasis based on the Integrated Disease Surveillance and response guidelines.^22^ It includes data on both clinically diagnosed and laboratory confirmed leishmaniasis cases, admissions, primary visceral leishmaniasis i.e. without history of visceral leishmaniasis treatment, relapse i.e. with history of visceral leishmaniasis treatment, post-kala-azar dermal leishmaniasis, a complication of visceral leishmaniasis and leishmaniasis HIV co-infection, and deaths. Before 2020, all forms of leishmaniasis (cutaneous, visceral and mucosal) were reported jointly in the monthly outpatient and in-patient registers. In 2020, an in-patient tool that collects primary visceral leishmaniasis, relapse and leishmaniasis HIV co-infection was introduced. In this study, we utilized aggregate data of on leishmaniasis from monthly outpatient reports (Health Management Information System forms [HMIS] 105) 2015–2022, inpatient monthly reports (HMIS 108) 2015–2022, and (HMIS 127b) 2020–2022.

### Study variables, data management, and analysis

We downloaded the data regarding clinically-diagnosed and laboratory-confirmed leishmaniasis cases and admissions, primary visceral leishmaniasis, relapse visceral leishmaniasis, post-kala-azar dermal leishmaniasis, and visceral leishmaniasis HIV co-infection, and death.in excel and imported in STATA version 16 software (StataCorp, College Station, Texas, USA) for analysis. Using data on leishmaniasis and visceral leishmaniasis cases, we calculated the annual prevalence of leishmaniasis as laboratory confirmed leishmaniasis cases divided by the population in the region. From 2020 to 2022, we assessed the proportions of primary, relapse, post-kala-azar dermal leishmaniasis, and HIV co-infection among admitted Visceral Leishmaniasis cases.

Quarterly prevalence of Visceral Leishmaniasis was calculated as the ratio of primary and relapse cases to the population, obtained from Uganda Bureau of Statistics.

Leishmaniasis admissions and hospitalization prevalence were determined using leishmaniasis cases as denominators. Case fatality rates were computed based on the proportion of deceased leishmaniasis cases. Line graphs were used to show trends, and Mann-Kendall test to assess the significance of trends. We used choropleth maps created with QGIS software to illustrate the spatial distribution of leishmaniasis in the region.

### Ethical Considerations

Our study utilized routinely generated aggregated surveillance data with no personal identifiers in health facility outpatient and in-patient monthly reports, obtained from the DHIS-2. The Uganda Public Health Fellowship Program is part of the National Rapid Response Team, and has been granted permission to access and analyse surveillance data in the DHIS-2 and other data such as survey and field investigation data to inform decision making in the control and prevention of outbreaks and public health programming. Additionally, the Ministry of Health (MoH) has also granted the program permission to disseminate the information through scientific publications. We stored the abstracted data set in a password-protected computer and only shared it with the investigation team. In addition, the Office of the Associate Director for Science, U.S. Centers for Disease Control and Prevention, determined that this study was not a human subjects research with the primary intent of improving use of surveillance data to guide public health planning and practice.

This activity was reviewed by CDC and was conducted consistent with applicable federal law and CDC policy.§ §See e.g., 45 C.F.R. part 46, 21 C.F.R. part 56; 42 U.S.C. §241(d); 5 U.S.C. §552a; 44 U.S.C. §3501 et seq.

## Results

### Trends in the prevalence of leishmaniasis, Karamoja Region, Uganda, 2015-2022

Reporting rates for the Karamoja region ranged from 96% to 100% during the review period (from 2015 to 2022). Overall, 4,008 clinically-diagnosed leishmaniasis cases were reported. Of these, 484 (11%) were confirmed. The average annual prevalence of leishmaniasis was 4 cases per 1,000,000 population in the Karamoja Region reported from 2015 to 2022. No trend in leishmaniasis prevalence was observed, Kendall’s score=-13, p=0.173 (Figure 1).

**Figure 1.**
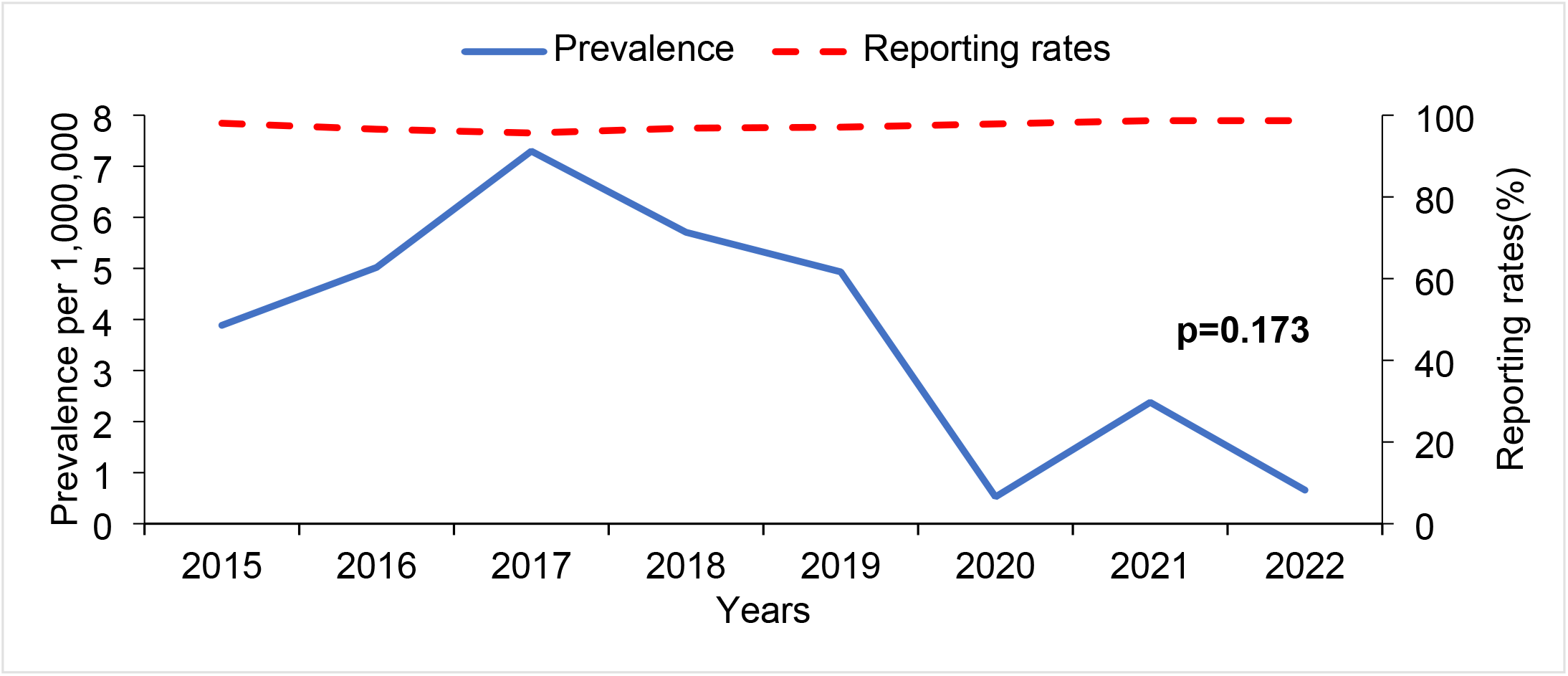
Trends in annual leishmaniasis prevalence, Karamoja Region, Uganda, 2015–2022

### Trends of the quarterly prevalence of visceral leishmaniasis, Karamoja Region, Uganda, 2020–2022

Over the review period, 602 visceral leishmaniasis cases were reported. Of these 540 (90%) were primary visceral leishmaniasis cases while 62 (10%) were relapse cases. Of the total cases, 4 (1%) were visceral leishmaniasis HIV-coinfected and 5 (1%) were post-kala-azar dermal leishmaniasis. The average quarterly prevalence of visceral leishmaniasis cases was 3 cases per million with the highest prevalence (5 cases per 1,000,000 population) reported in the quarter 3 of 2022 (Figure 2). From 2020 to 2022, there was an increasing trend in the quarterly prevalence of visceral leishmaniasis, Kendall’s score=36, p=0.016.

**Figure 2.**
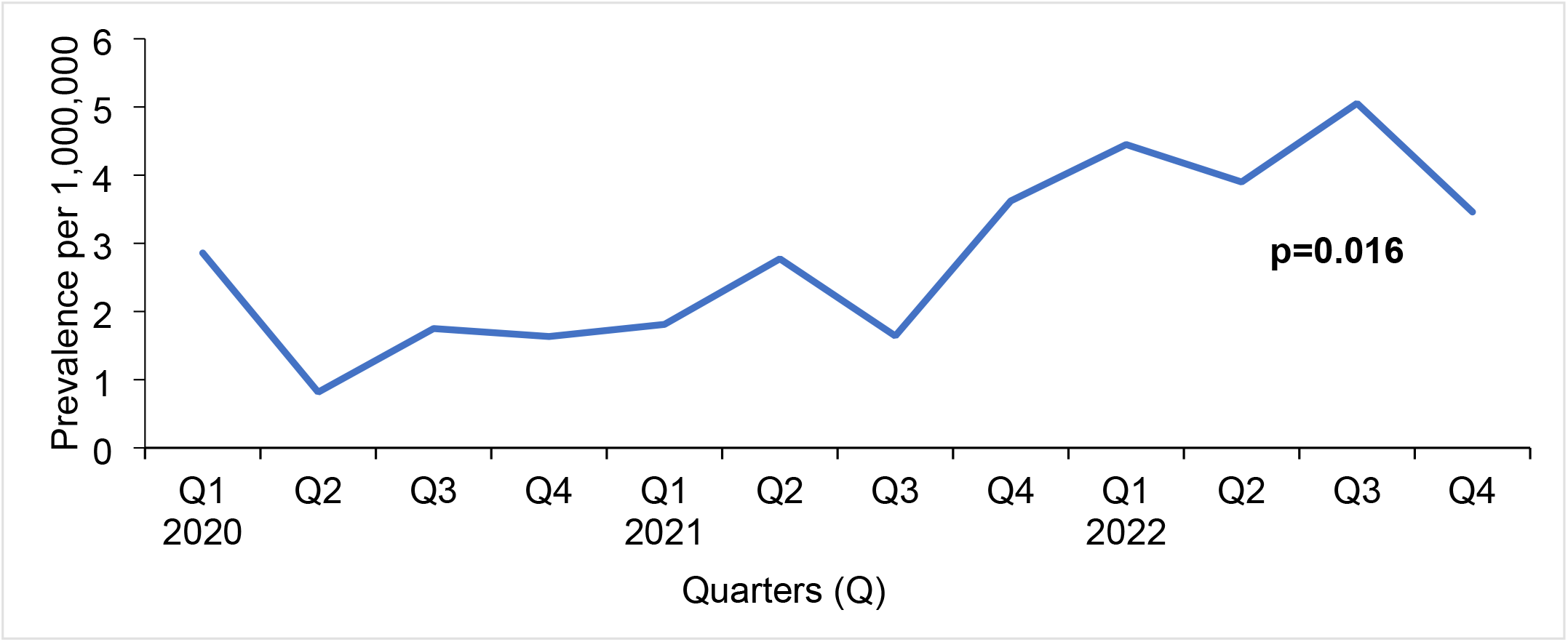
Trends in quarterly prevalence of visceral leishmaniasis, Karamoja Region, Uganda, 2020–2022.

### Trends of leishmaniasis admissions, Karamoja Region, Uganda, 2015–2022

From 2015 to 2022, the average annual prevalence of leishmaniasis admission in the Karamoja region was 52 cases per 100 cases. The highest incidence of leishmaniasis admissions were reported in the years 2020 (87 leishmaniasis admissions per 100 cases), 2021 (93 leishmaniasis admissions per 100 cases), and 2022 (92 leishmaniasis admissions per 100 cases) (Figure 3). Overall there was an increasing trend of leishmaniasis admissions, Kendall’s score= 23, p=0.006.

**Figure 3.**
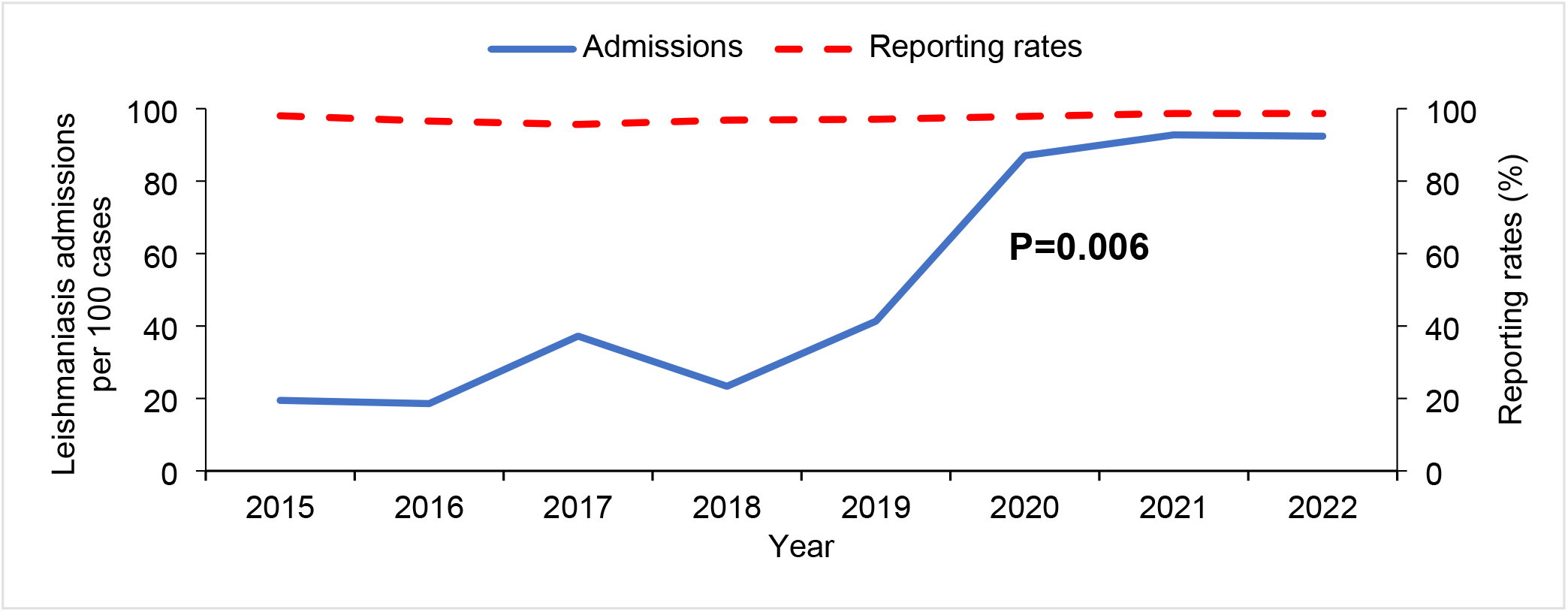
Trends in hospital admissions due to leishmaniasis in Karamoja Region, Uganda, 2015–2022.

### Leishmaniasis case fatality rate, Karamoja region, Uganda, 2015–2022

During the review period, the average annual CFR was 5% (Range: 0-11%), with no deaths reported in 2018, 2019, and 2021.The highest case fatality rates was recorded in 2020. No trends were observed in the case fatality rate, Kendall’s score= -2, p=0.947 (Figure 4).

**Figure 4.**
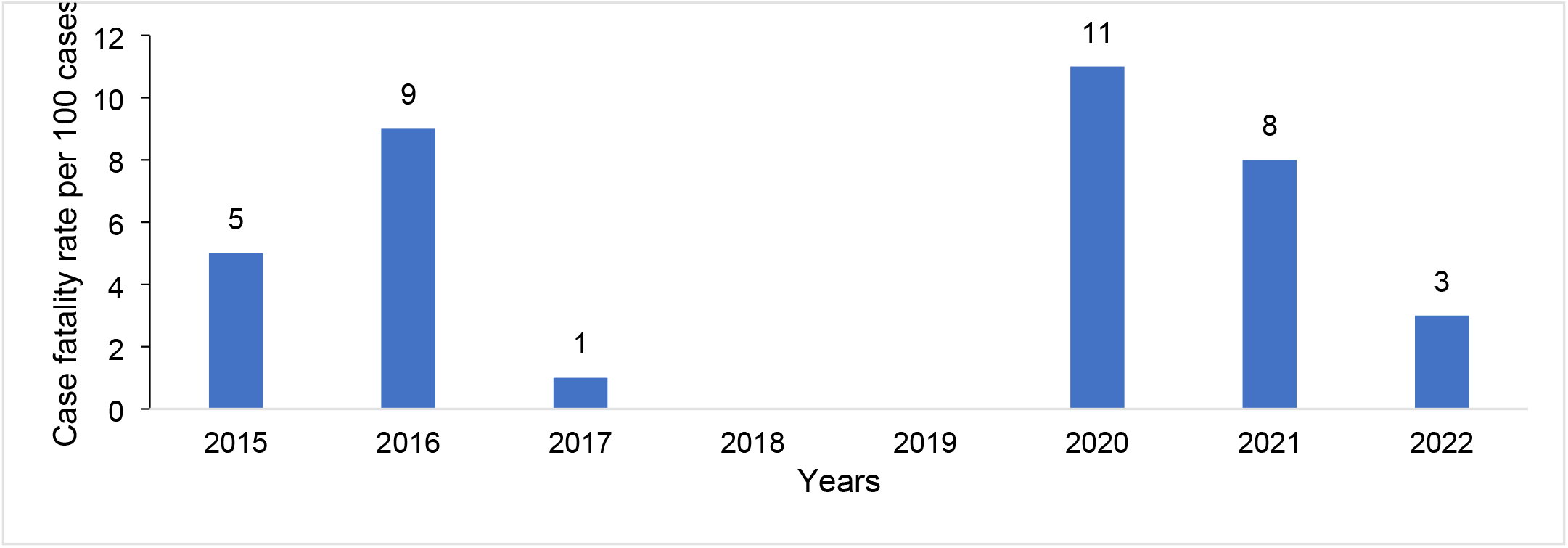
Trends in leishmaniasis case fatality rate, Karamoja Region, Uganda, 2015–2022

### Spatial distribution of Leishmaniasis, Karamoja Region, Uganda, 2015–2022

Within the Karamoja Region, Amudat District reported the highest average annual prevalence of both leishmaniasis (477 per 1,000,000 population) and visceral leishmaniasis (139 per 100,000 population). Abim, Kaabong, Karenga, and Nabilatuk did not report both leishmaniasis and visceral leishmaniasis cases during the reporting period.

**Figure 5.**
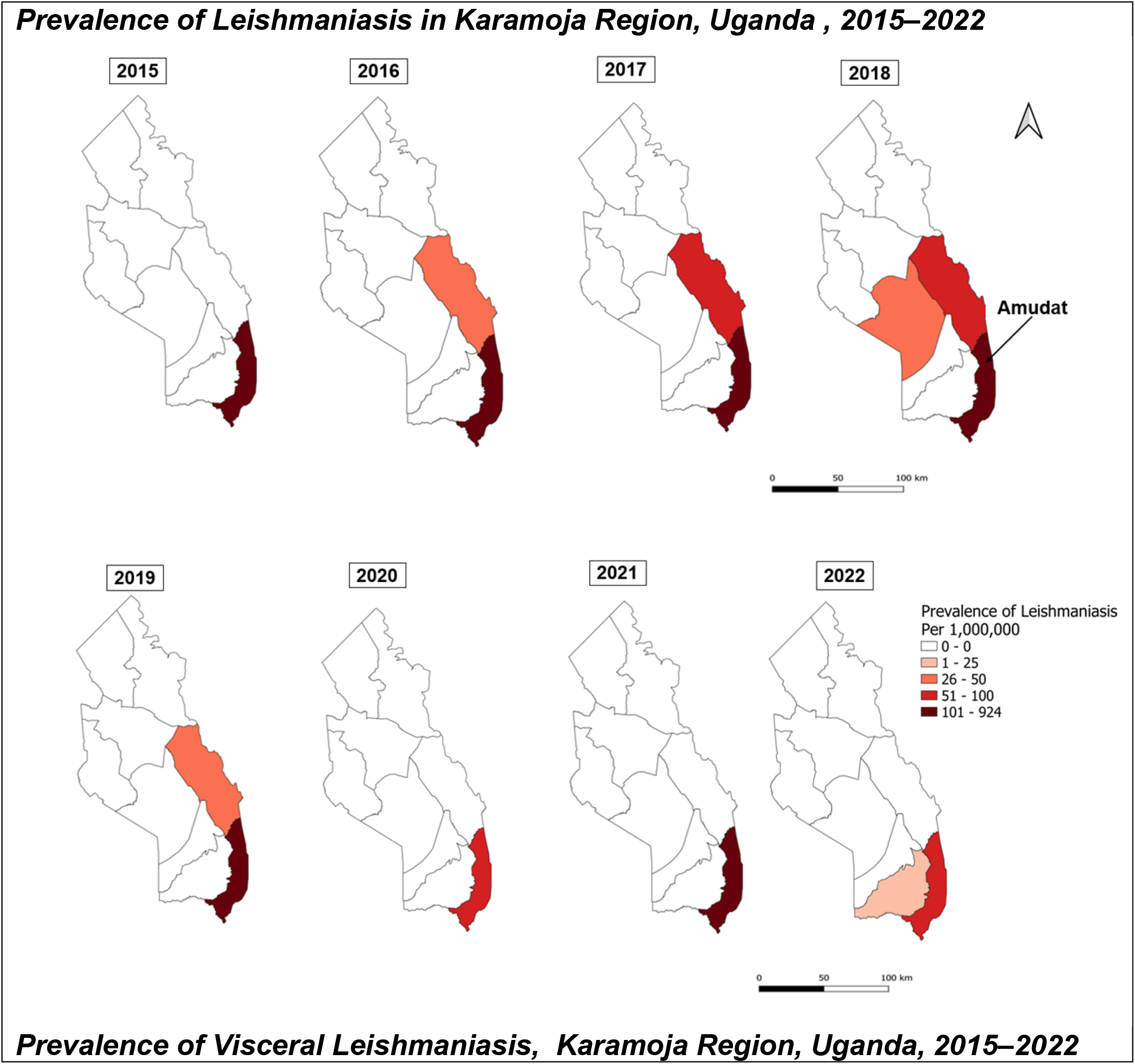
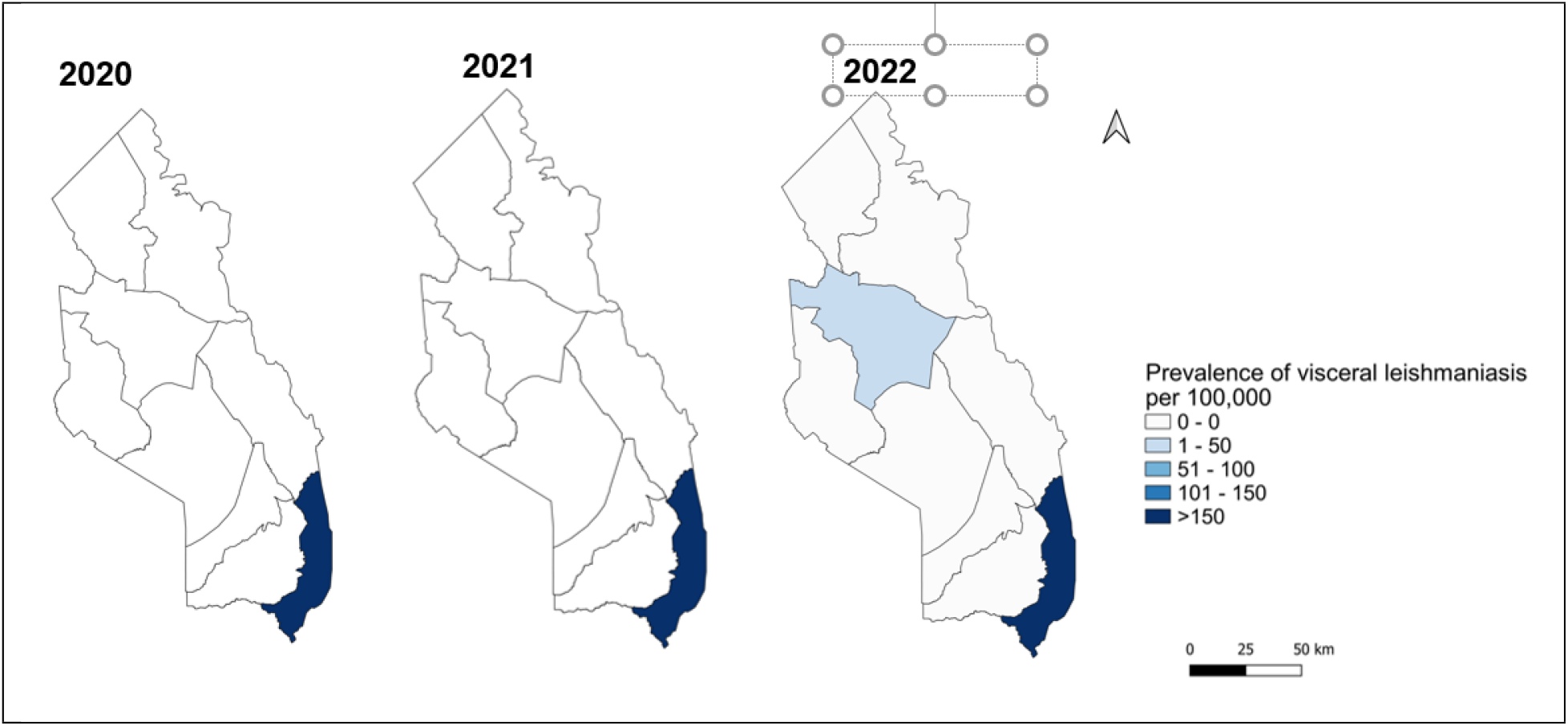
Spatial distribution of leishmaniasis and visceral leishmaniasis, Karamoja Region, Uganda, 2015–2022

## Discussion

In this analysis to examine the temporal trends and spatial distribution of leishmaniasis in an endemic Karamoja Region of Uganda, 2015–2022, only 11% of the clinically-diagnosed leishmaniasis cases were laboratory confirmed. Over the review period, no changes were observed in the annual prevalence of leishmaniasis. While most visceral leishmaniasis cases were primary, we identified relapse, post-kala-azar dermal leishmaniasis, and visceral leishmaniasis HIV coinfected cases between 2020 and 2022. A rising trend in quarterly prevalence of visceral leishmaniasis occurred from quarter one (Q1) 2020 to quarter four (Q4) 2022, along with an increase in hospital admissions due to leishmaniasis. The average annual CFR remained <1%, with no deaths reported in 2018, 2019, and 2021. Amudat District had the highest leishmaniasis prevalence.

We identified gaps in case detection, with only 11% of clinically-diagnosed cases confirmed by laboratory investigations. Leishmaniasis shares clinical features with common regional illnesses like malaria, typhoid, and tuberculosis, emphasizing the importance of laboratory confirmation in case detection^23^. In the East African region, numerous factors may hinder laboratory confirmation for leishmaniasis including low proportions of health facilities with microscopes and microscopy supplies and trained staff in leishmaniasis testing^24^. Such existing gaps may hinder achievement of leishmaniasis targets of 85% of all leishmaniasis cases detected.

In contrast to the global declining trends in leishmaniasis prevalence, no significant trends were observed in the Karamoja Region for leishmaniasis cases from 2015 to 2022^25,26^. The Karamoja Region like the rest of East Africa faces unique challenges that may prevent declines despite targets. The Karamoja Region like the rest of East Africa faces unique challenges that may prevent declines despite targets, including poor at-risk population with limited access to healthcare and insecurity in endemic regions^19^.

Our findings further indicated an increase in the quarterly prevalence of visceral leishmaniasis cases from 2020 to 2022.This trend is similar to global leishmaniasis surveillance data that indicated an increase in visceral leishmaniasis in the East African Region during 2020^11^. During this period, vital control programs supporting the region, such as the Accelerating the Sustainable Control and Elimination of Neglected Tropical Diseases, were terminated. This resulted in the scaling down of control efforts^27^. The increase in visceral leishmaniasis cases following termination of control programmes may undo previous gains towards achievement of elimination targets. Furthermore, we found Visceral Leishmaniasis relapse and post-kala-azar dermal leishmaniasis (PKDL) cases. The presence of these cases contributes to the maintenance of infectious reservoirs, posing a threat to the achievement of elimination targets^28^.

Our findings revealed a considerable number of hospital admissions due to leishmaniasis. This suggests a sustained burden of visceral leishmaniasis whose illness and management often requires hospitalisation^29^. Admissions due to leishmaniasis impose a substantial economic burden on households, with a median expenditure of 450 US dollars per hospital stay due to leishmaniasis^30^. We found that hospital admissions linked to leishmaniasis increased. This might suggest gaps in treatment of leishmaniasis resulting in leishmaniasis complications that may require in-patient admissions.

The average annual CFR was still greater than the 1% target by 2030. This indicated that Uganda is yet to achieve the control leishmaniasis and eliminate visceral leishmaniasis as a public health elimination. The no trend observed in CFR and its substantial variation in the CFR from 0 to 11% suggests a need for intensifying efforts to ensure targets are met. Such efforts include improving prompt diagnosis of visceral leishmaniasis using rapid diagnostic kits and increasing access to prompt effective treatment.

In regards to the current distribution, Amudat had the highest prevalence of both leishmaniasis and visceral leishmaniasis. This pattern may be attributed to presence of a referral leishmaniasis treatment centre at Amudat Hospital^31^.

Our findings should be interpreted with considerations of the following limitations. Our use of facility-based data may lead to an underestimation of the true disease burden, capturing only individuals seeking healthcare. Previous studies have indicated that surveillance data may underreport the burden of leishmaniasis up to 8 fold for visceral leishmaniasis and 5 fold for cutaneous leishmaniasis^9^. Additionally, our reliance on aggregate data restricted further exploration of prevalence in population subcategories and specific clinical types of leishmaniasis.

## Conclusion

The current burden of leishmaniasis coupled with gaps in case detection threatens the achievement of control of leishmaniasis targets. While the increasing trends of visceral leishmaniasis and variation in case fatality rates question whether achieved visceral leishmaniasis targets will be met by 2030. Existing interventions should be intensified to including vector control, rapid diagnostic kits for early detection and prompt treatment should be intensified to sustain the achievement of elimination targets.

## Data Availability

The datasets upon which our findings are based belong to the Uganda Public Health Fellowship Program. The data sets can be availed upon reasonable request from the corresponding author with permission from the Uganda Public Health Fellowship Program.

## List of abbreviations

CFR: Case Fatality Rate

DHIS-2: District Health Information System 2

HMIS: Health Management Information System

NTD: Neglected Tropical Disease

PKDL: Post-kala-azar Dermal Leishmanisis

## Acknowledgements

We would like to thank the Ministry of Health for providing access to DHIS2 data that was used for this analysis. We appreciate the technical support provided by the Division of Surveillance, Information and Knowledge Management and the Division of Vector Control. Finally, we thank the US-CDC for supporting the activities of the Uganda Public Health Fellowship Program.

